# Predicting dengue in the Philippines using artificial neural network

**DOI:** 10.1101/2020.10.08.20209718

**Authors:** Bryan Zafra

**Affiliations:** College of Medicine, University of the City of Manila, Manila, Philippines

**Keywords:** dengue, Philippines, artificial neural network, climate change

## Abstract

Dengue fever is an infectious disease caused by *Flavivirus* transmitted by *Aedes* mosquito. This disease predominantly occurs in the tropical and subtropical regions. With no specific treatment, the most effective way to prevent dengue is vector control. The dependence of Aedes mosquito population on meteorological variables make prediction of dengue infection possible using conventional statistical and epidemiologic models. However, with increasing average global temperature, the predictability of these models may be lessened employing the need for artificial neural network. This study uses artificial neural network to predict dengue incidence in the entire Philippines with humidity, rainfall, and temperature as independent variables. All generated predictive models have mean squared logarithmic error of less than 0.04.

## 1. Introduction

Dengue fever is an infectious disease caused by *Flavivirus* transmitted by the vector mosquito *Aedes*. The dengue virus has 4 serotypes (DENV-1, DENV-2, DENV-3, DENV-4) which mostly occur in the urban and sub-urban areas in tropical and subtropical regions. It is estimated that 50 million people have dengue infection globally per annum. In the Philippines, an estimated 170,000 cases occurred annually. Currently, there are no specific treatments for dengue and the most effective way to prevent it is through vector control. [1-6]

The 2 most important vectors for dengue transmission are: Aedes aegypti and Aedes albopictus. The lifecycle of these vectors is divided into egg, larva, pupa, and adult stages; which is heavily influenced by different meteorological, geological, and anthropological variables. The typical adult mosquito lays egg just above the waterline of a stagnant water. It will take 48 hours in warm, humid environment for the embryo to develop. Once developed, the eggs are tolerant to desiccation up to more than a year.[1] The adult mosquito then typically emerges after 10 days. The adult female mates and takes blood meal necessary for egg maturation. The blood meal biting activity takes place in the morning and afternoon. [7, 8] However, blood meals also do occur at night in lighted rooms. [3]

Temperature, rainfall, and relative humidity affects the transmission of dengue. [9] Annual rainfall of more than 200 cm provides conducive growth of Aedes population. [10] Mosquito population growth is more abundant in sea level up to 500 meters above sea level, although they can thrive up to 1,200 meters. [11] Climate variability strongly influence dengue epidemic. Maximal temperature of more than 32°C, maximal relative humidity of more than 95% influence the incubation period, feeding frequency, and longevity of Aedes. [12] There is low mosquito mortality in temperature between 15°C to 30°C. Pupae development occurs in less than 1 day in 32°C to 34°C but takes 4 days in 22°C. [13-17]

With the dependence of Aedes mosquito population dynamics on weather variables; climate change, undoubtedly, will have its impact on the spread of dengue infection. It is estimated that the average global temperature will increase by 2°C to 4.5°C by year 2100. [18] There are many researches that predicted dengue infection using weather variables. Different statistical models were employed such as: Poisson regression, autoregressive integrated moving average (ARIMA), seasonal autoregressive integrated moving average (SARIMA) [19, 22], negative binomial, quasi-likelihood regression [19, 20], distributed lag non-linear model (DLNM) [21]. There are also attempts to employ machine learning techniques such as random forest and gradient boosting to make dengue infection predictions. [23-25] These predictive models have varying degree of predictions on dengue incidence.

With the ease of accessibility and less expensive computing power available nowadays, there is increased application of artificial neural network in making predictions in different areas of science. Coupled with the use of programming and software packages such as Python and TensorFlow, this research will attempt to predict dengue incidence in the Philippines using artificial neural network.

## 2. Materials and Methods

### 2.1. Study Setting

The study was conducted in all 17 administrative regions in the Philippines: Region 1 to 12 (including 4-A and 4-B), Autonomous Region of Muslim Mindanao (ARMM), Cordillera Autonomous Region (CAR), and CARAGA. The Philippines has two seasons: wet (June to October) and dry (November to May). However, there are 4 climate types in the Philippines based on modified Coronas classification: Type I (dry from November to April, we from May to October), Type II (seasonal rainfall from November to December), Type III (same as Type I but with maximum rainfall from May to October), and Type IV (evenly distributed rainfall annually). [26]

### 2.2. Data Collection

Data on dengue incidence is freely available in the Department of Health (DOH) website as PDF reports. These dengue reports are sum of all case definition of a dengue case with or without the confirmation of polymerase chain reaction. This is attributed to the limited resources available especially in remote and rural areas where dengue case definition is based on signs and symptoms only. The meteorological data: humidity (as %), rainfall (as mm), temperature (as °C) were requested from the Philippine Atmospheric, Geophysical and Astronomical Services Administration (PAGASA) and were received as excel files. Missing dengue values were addressed by getting the average value from the same region from different year period but of the same week time frame. Missing meteorological values were filled using the average value from same weather station from different year period but of the same month time frame. These files from DOH and PAGASA were encoded to CSV files for data analysis. Dengue data were reported on a weekly basis from each administrative regions while the meteorological data were reported on a monthly basis from each weather stations where most administrative regions have 2 to 3 weather stations. To reconcile these differences, the weekly tally of dengue were summed up to reflect a month value while the meteorological values were averaged out from the weather stations to reflect the regional value. The data on dengue and weather variables is from year 2013 to 2018.

### 2.3. Artificial Neural Network

Data were analyzed in Python 3 using Jupyter Notebook as the interface while also employing several libraries such as Numpy, Pandas, Keras, and Tensorflow. A training and test set were created for each region. The training set consists of data from 2013 to 2017 and the test set contains data from 2018. An artificial neural network was created with 1 input layer, 6 hidden layers, and 1 output layer as shown in Figure 1. The input layer is composed of humidity, rainfall, and temperature. The output layer is composed of dengue incidence.

**Figure 1.**
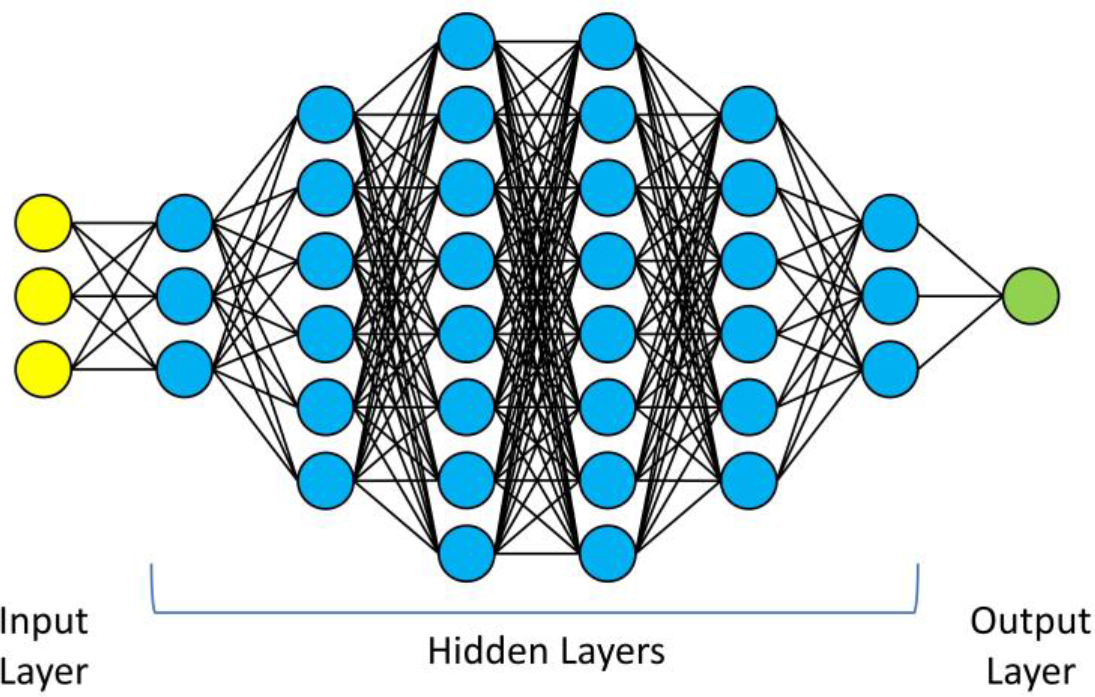
Artificial neural network architecture.

The artificial neural network uses rectified linear unit (*ReLu*) as the activation function and adaptive moment estimation (*Adam*) as an optimiser. The artificial neural network was trained with batch size of 24 in 500 epochs for each administrative region. The created model from the training set was used to predict the dengue values in the test set. The prediction was evaluated using mean squared logarithmic error (MSLE).

## 3. Results

Table 1 shows the descriptive statistics of monthly humidity, rainfall, temperature, and dengue incidence for each administrative region. ARMM has the lowest average dengue incidence of 157.26 while Region 4-A has the highest at 2,616.97. NCR has the lowest average humidity of 75.24% while CAR has the highest at 87.25%. Region 12 has the lowest average rainfall of 81.99 mm, while Region 7 has the highest at 1,130.61 mm. CAR has the lowest average temperature of 19.47°C, while Region 11 has the highest at 28.7°C.

**Table 1.** Descriptive statistics of monthly humidity, rainfall, temperature, and dengue incidence in each administrative region of the Philippines (2013 to 2018).

| Region | Humidity (%) |  |  |  | Rainfall (mm) |  |  |  | Temperature (°C) |  |  |  | Dengue Incidence |  |  |  |
| --- | --- | --- | --- | --- | --- | --- | --- | --- | --- | --- | --- | --- | --- | --- | --- | --- |
|  | Mean | SD | Min | Max | Mean | SD | Min | Max | Mean | SD | Min | Max | Mean | SD | Min | Max |
| ARMM | 76 | 2.69 | 68 | 80 | 160.07 | 90.76 | 2.8 | 393.4 | 28.22 | 0.66 | 26.8 | 29.9 | 157.26 | 80.76 | 16 | 495 |
| CAR | 87.25 | 4.12 | 78 | 96 | 307.52 | 400.32 | 0 | 1822.6 | 19.47 | 1.01 | 15.8 | 21.4 | 681.99 | 670.35 | 58 | 2965 |
| CARAGA | 82.74 | 3.11 | 76 | 89.33 | 298.01 | 200.62 | 47.4 | 1126.47 | 28.10 | 1.02 | 24.97 | 30.5 | 812.79 | 506.97 | 144 | 2574 |
| NCR | 75.24 | 7.16 | 61.33 | 89 | 195.18 | 201.78 | 0.1 | 890.6 | 28.45 | 1.26 | 25.03 | 30.97 | 1569.15 | 1329.74 | 78 | 7977 |
| 1 | 79.41 | 4.63 | 71.33 | 88.67 | 198.22 | 274.02 | 0 | 1069.93 | 27.61 | 1.22 | 23.87 | 29.9 | 1024.88 | 1013.94 | 99 | 4917.99 |
| 2 | 84.36 | 2.27 | 79.8 | 88.82 | 171.85 | 125.91 | 8.56 | 526.84 | 26.31 | 2.29 | 21.62 | 29.7 | 697.51 | 786.54 | 57 | 3821 |
| 3 | 79.70 | 3.93 | 71.67 | 87.83 | 249.67 | 189.07 | 23.72 | 803.02 | 27.75 | 1.21 | 24.58 | 30.27 | 2515.51 | 2202.24 | 139.98 | 13134 |
| 4-A | 83.92 | 2.62 | 77 | 88.33 | 237.88 | 154.1 | 13.08 | 540.5 | 26.91 | 1.26 | 23.82 | 29.32 | 2616.97 | 2280.62 | 197.9 | 14568.15 |
| 4-B | 81.37 | 3.17 | 73.67 | 86.83 | 183.92 | 139.7 | 12.08 | 599.25 | 27.93 | 0.75 | 26.30 | 29.82 | 769.62 | 580.72 | 102.5 | 3298.83 |
| 5 | 84.67 | 2.02 | 81 | 88.5 | 248.7 | 163.84 | 23.35 | 733.18 | 27.71 | 1.06 | 25.15 | 29.58 | 234.18 | 130.55 | 36 | 500 |
| 6 | 81.03 | 2.15 | 76 | 86.48 | 176.2 | 120.26 | 3 | 476.5 | 28.42 | 0.8 | 26.2 | 30.2 | 1424.78 | 1391.65 | 147 | 6924 |
| 7 | 81.52 | 2.75 | 75 | 86.09 | 1130.61 | 73.88 | 1.53 | 271.23 | 28.09 | 0.8 | 25.9 | 29.73 | 1687.05 | 1291.86 | 102 | 5307.99 |
| 8 | 85.02 | 2.56 | 78.17 | 90.53 | 297.37 | 199.86 | 33.17 | 1045.8 | 27.74 | 0.89 | 25.41 | 29.27 | 442.98 | 298.53 | 31.02 | 1840.02 |
| 9 | 81.66 | 2.32 | 75 | 86 | 163.37 | 103.66 | 2.65 | 522.2 | 28.3 | 0.47 | 26.6 | 29.3 | 518.76 | 216.22 | 184 | 1155 |
| 10 | 84.73 | 2.66 | 77.5 | 89.5 | 180.46 | 103.38 | 2.9 | 443.95 | 26.11 | 0.68 | 24.3 | 27.65 | 1078.71 | 699.18 | 344 | 3414 |
| 11 | 78.7 | 3.36 | 70 | 85 | 161.47 | 94.14 | 14.2 | 430.5 | 28.7 | 0.69 | 26.5 | 30.6 | 599.91 | 388.52 | 116 | 2581 |
| 12 | 77.53 | 4.72 | 65 | 84 | 81.99 | 51.8 | 0.1 | 256.8 | 27.99 | 0.93 | 26.5 | 31.1 | 1019.04 | 573.58 | 278 | 3443 |

ARMM – Autonomous Region in Muslim Mindanao
CAR – Cordillera Autonomous Region
NCR – National Capital Region

Python code implementation of the artificial neural network in Figure 1.

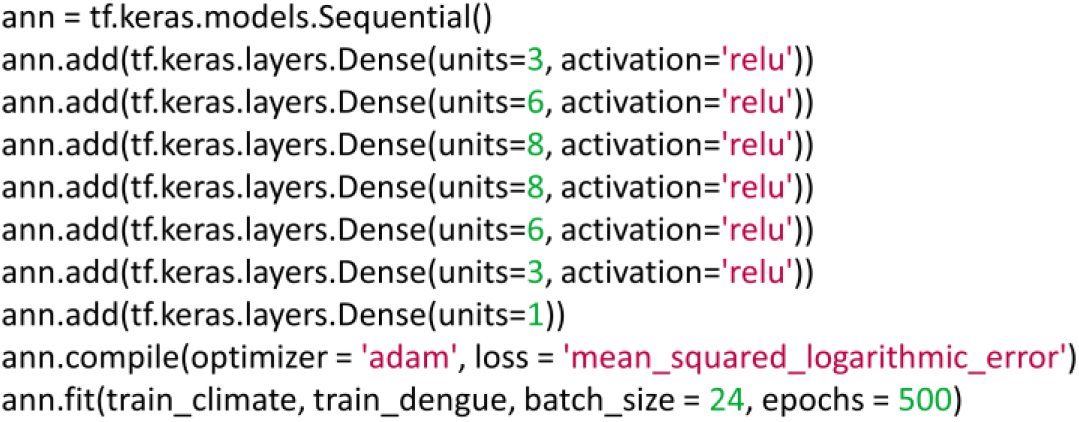

The resulting predictive models from the artificial neural network has a mean squared logarithmic error of less than 0.04 in all administrative regions. Table 2 provides the values of MSLE in each region. Region 9 has the lowest MSLE of 0.006 while Region 2 has the highest at 0.0376. Figure 2 provides comparison of the actual and predicted dengue incidence in each administrative regions using the generated predictive models.

**Table 2.** Mean squared logarithmic error of each administrative region in the Philippines.

| Region | MSLE |
| --- | --- |
| ARMM | 0.008775361 |
| CAR | 0.020651964 |
| CARAGA | 0.009911519 |
| NCR | 0.021502396 |
| Region 1 | 0.019390212 |
| Region 2 | 0.03755766 |
| Region 3 | 0.015200399 |
| Region 4-A | 0.007498101 |
| Region 4-B | 0.01333835 |
| Region 5 | 0.0097335195 |
| Region 6 | 0.009920411 |
| Region 7 | 0.014176335 |
| Region 8 | 0.014938806 |
| Region 9 | 0.006088777 |
| Region 10 | 0.014615116 |
| Region 11 | 0.012485982 |
| Region 12 | 0.014623637 |

**Figure 2.**
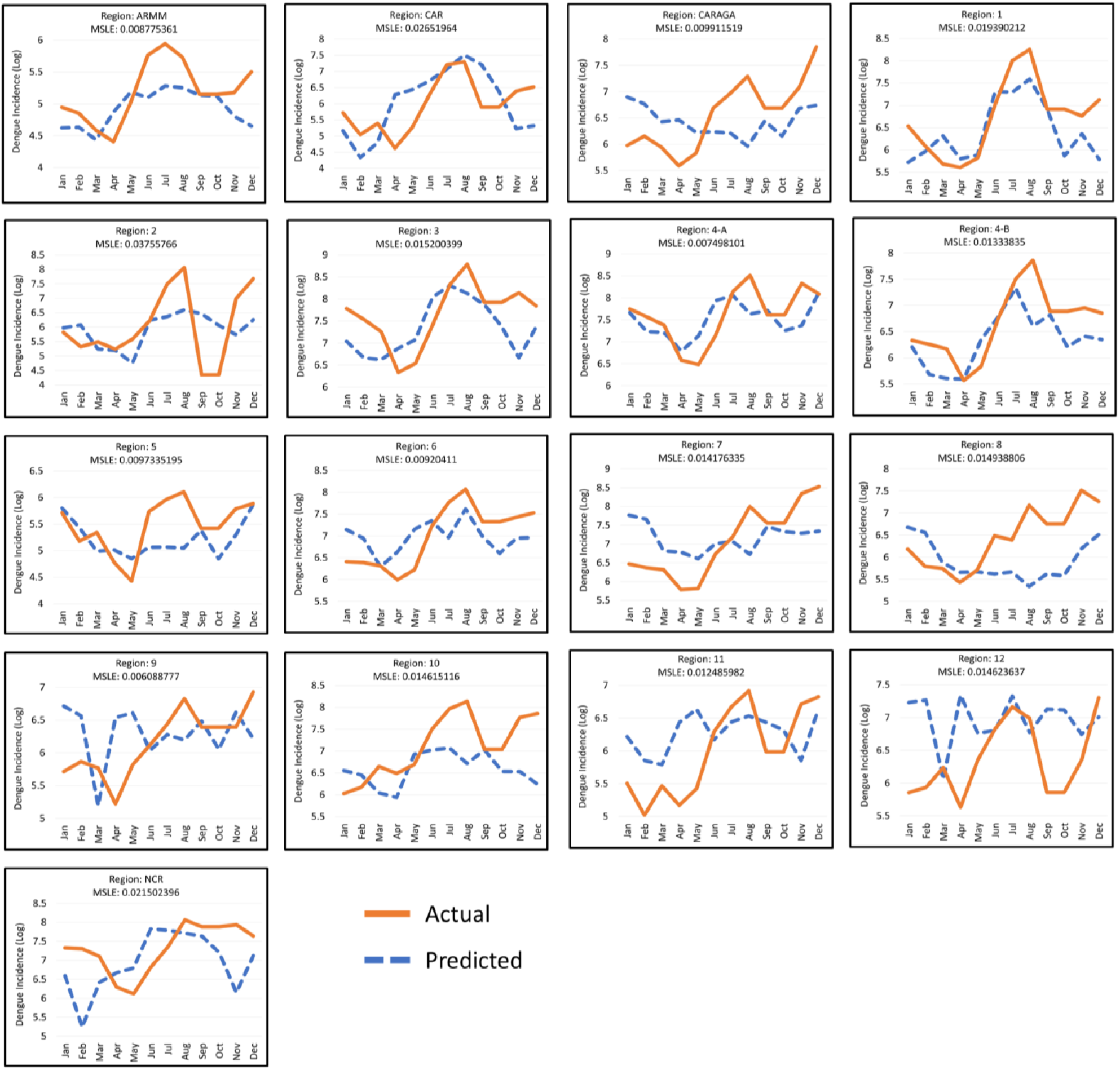
Comparison of the actual and predicted dengue incidence in all administrative regions in the Philippines (2018).

## 4. Discussion

Although there is low MSLE in each administrative region, visual inspection of the actual and predicted dengue incidence revealed that there are predictive models that are better than the other. It should be noted that the predictive models are unique to each region since they are trained separately and have their own artificial neural network even though they have the same network architecture. The administrative regions that have visually performed well are: ARMM, CAR, Region 1, 3, 4-A, and 4-B. The predictive models also had inefficiencies in identifying the peak dengue incidence which are evident in Region 2, 5, 10, and ARMM. The worst predictive models are in CARAGA, NCR, Region 2, 7, 8, 9, 11, and 12.

Predictions made by an artificial neural network is different from statistical or epidemiological modelling. Artificial neural network utilizes collection of artificial neurons that take the weighted inputs, pass it through an activation function, to produce an output. [27] In this study, there are 3 inputs: humidity, rainfall, and temperature; and there is 1 output which is a single value of dengue incidence. The idea is, at any given values of the 3 input variables, what will be the resulting dengue incidence. The rectified linear unit (ReLU) [28] is the activation function used to avoid having negative values for the dengue incidence. These multiple units and layers of computation can make better predictions.

There are several limitations that was encountered in this study. The study encompasses entire administrative region, which means micro-climate variability from each city or municipality can be a contributing factor to the dengue incidence. The meteorological variables provided by PAGASA was limited to 3 (humidity, rainfall, temperature), although the request includes flood occurrence and average sunlight. These 3 significant meteorological variables appeared in the researches done in the Philippines. [23, 29] However, flood occurrence may help in dengue incidence prediction because flushing can potentially reduce dengue incidence. [30] Average sunlight have inconclusive relationship to dengue incidence using statistical model. [31] However, this might be proven otherwise if artificial neural network is use.

The impact of climate change can influence the transmission of dengue to other places other than the tropical and subtropical regions. By the end of this century, dengue epidemic potential for Aedes aegypti could occur in 10 European cities (Madeira, Malaga, Athens, Rome, Nice, Paris, London, Amsterdam, Berlin, Stockholm) with continued current rate of greenhouse gas emission. [32] The complexities of weather and climate influences on dengue transmission are not easily modelled with statistical approach [33] which makes artificial neural network more helpful in predicting dengue incidence more accurately.

## 5. Conclusion

Close fidelity on the predicted and actual dengue incidence in some administrative region in the Philippines prove that artificial neural network can be implemented for predicting dengue. Further work can be done in optimizing the artificial neural network architecture: number of neurons, number of hidden layers, and additional input meteorological variables (flood occurrence, average sunlight). It is recommended that future research be focused on the city or municipal level of dengue cases and weather variables measurement to be able to create a local public health policy.

## Data Availability

The dengue data is available in the Department of Health website. The weather variables are requested from Philippine Atmospheric, Geophysical, and Astronomical Services in the Philippines.

## References

[1] World Health Organization. Comprehensive Guidelines for Prevention and Control of Dengue and Dengue Haemorrhagic Fever (Revised and expanded edition). 2011. https://apps.who.int/iris/handle/10665/204894 [Accessed July 2020].

[2] Undurraga E, Edillo F, Erasmo JN, Alera MT, Yoon IK, Largo F, Shepard D. “Disease Burden of Dengue in the Philippines: Adjusting for Underreporting by Comparing Active and Passive Dengue Surveillance in Punta Princesa, Cebu City,” Am J Trop Med Hyg, 2017 Apr 5; 96(4): 887–898.

[3] Gubler DJ. “Dengue and dengue hemorrhagic fever,” Clin Microbiol Rev, 1998; 11(3):480–496.

[4] Gubler DJ. “Epidemic dengue/dengue hemorrhagic fever as a public health, social and economic problem in the 21st century,” Trends Microbiol, 2002; 10(2):100–103.

[5] Rigau-Pérez JG, Clark GG, Gubler DJ, Reiter P, Sanders EJ, Vorndam AV. Dengue and dengue haemorrhagic fever, Lancet. 1998; 352(9132):971–977.

[6] Singh M, Chakraborty A, Kumar S, Kumar A. “The epidemiology of dengue viral infection in developing countries: A systematic review,” J Health Res Rev, 2017; 4(3):104–107.

[7] Nelson MJ, Self LS, Pant CP, Slim U. “Diurnal periodicity of attraction to human bait of Aedes aegypti in Jakarta, Indonesia,” J Med Entomol, 1978; 14: 504–10.

[8] Sheppard PM, Maedonald WW, Tonk RJ, Grab B. “The dynamics of an adult population of Aedes aegypti in relation to DHF in Bangkok,” J Animal Ecology, 1969; 38: 661-2.

[9] Naish S, Dale P, Mackenzie JS, McBride J, Mengersen K, Tong S. “Climate change and dengue: a critical and systematic review of quantitative modelling approaches,” BMC Infect Dis, 2014; 14(1):167.

[10] Ehelepola NDB, Ariyaratne K, Buddhadasa WMNP, Ratnayake S, Wickramasinghe M, Promprou S, et al. “A study of the correlation between dengue and weather in Kandy City, Sri Lanka (2003 -2012) and lessons learned,” Infect Dis Poverty, 2015;4:42.

[11] Kalra NL, Kaul SM, Rastogi RM. “Prevalence of Aedes aegypti and Aedes albopictus vectors of DF/DHF in North, North-East and Central India,” Dengue Bulletin, 1997; 21: 84–92.

[12] Descloux E, et al. “Climate-based models for understanding and forecasting dengue epidemics,” PLoS Negl Trop Dis, 2012; 6(2): e1470.

[13] Focks DA, Haile DG, Daniels E, Mount GA. “Dynamic life table model for Aedes aegypti (Diptera: Culicidae): analysis of the literature and model development,” J Med Entomol, 1993; 30: 1003-1017.

[14] Focks DA, Haile DG, Daniels E, Mount GA. “Dynamic life table model for Aedes aegypti (Diptera: Culicidae): simulation results and validation,” J Med Entomol, 1993; 30: 1018–1028.

[15] Focks DA, Haile DG, Daniels E, Keesling JE. “A simulation model of the epidemiology of urban dengue fever: literature analysis, model development, preliminary validation, and samples of simulation results,” Am J Trop Med Hyg, 1995; 53: 489–506.

[16] Hopp MJ, Foley JA. “Global-scale relationships between climate and the Dengue fever vector, Aedes aegypti,” Clim Change, 2001; 48: 441–463.

[17] Focks DA, Brenner RJ, Hayes J, Daniels E. “Transmission thresholds for for dengue in terms of Aedes aegypti pupae per person with discussion of their utility in source reduction efforts,” Am J Trop Med Hyg, 2002; 62: 11–18.

[18] Schnoor JL. “The IPCC fourth assessment,” Environ Sci Technol, 2007; 41: 1503.

[19] Chumpu R, Khamsemanan N, Nattee C. “The association between dengue incidences and provincial-level weather variables in Thailand from 2001 to 2014,” PLoS ONE, 2019; 14(12): e0226945.

[20] Choi Y, et al. “Effects of weather factors on dengue fever incidence and implications for interventions in Cambodia,” BMC Public Health, 2016; 16(1):241.

[21] Chuang T-W, Chaves LF, Chen P-J. “Effects of local and regional climatic fluctuations on dengue outbreaks in southern Taiwan,” PLoS ONE, 2017; 12(6): e0178698.

[22] Xuan LTT, Hau PV, Thu DT, Toan DTT. “Estimates of meteorological variability in association with dengue cases in a coastal city in northern Vietnam: an ecological study,” Glob Health Action, 2014, 7: 23119.

[23] Carvajal T, et al. “Machine learning methods reveal the temporal pattern of dengue incidence using meteorological factors in metropolitan Manila, Philippines,” BMC Infectious Diseases, (2018) 18:183.

[24] Jain R, Sontisirikit S, Iamsirithaworn S, Prendinger H. “Prediction of dengue outbreaks based on disease surveillance, meteorological and socio-economic data,” BMC Infectious Diseases, (2019) 19:272.

[25] Bakar AA, Kefli Z, Abdullah S, Sahani M. “Predictive models for dengue outbreak using multiple rulebase classifiers,” 2011 International Conference on Electrical Engineering and Informatics (ICEEI), Bandung, Indonesia, 17–19 July 2011. DOI: 10.1109/ICEEI.2011.6021830.

[26] Seposo XT, Dang TN, Honda Y. “Exploring the effects of high temperature on mortality in four cities in the Philippines using various heat wave definitions in different mortality subgroups,” Glob Health Action, 2017; 10:1368969.

[27] Alzahrani R, Parker A. “Neuromorphic Circuits With Neural Modulation Enhancing the Information Content of Neural Signaling,” International Conference on Neuromorphic Systems 2020 DOI: 10.1145/3407197.3407204.

[28] Nair V, Hinton G. “Rectified Linear Units Improve Restricted Boltzmann Machines”, 27th International Conference on International Conference on Machine Learning, ICML’10, USA: Omnipress, pp. 807–814.

[29] Iguchi J, Seposo X, Honda Y. “Meteorological factors affecting dengue incidence in Davao, Philippines,” BMC Public Health, (2018) 18:629.

[30] Benedum CM, Seidahmed OME, Eltahir EAB, Markuzon N. “Statistical modeling of the effect of rainfall flushing on dengue transmission in Singapore,” PLoS Negl Trop Dis, 2018; 12(12): e0006935.

[31] Shang C-S, et al. “The Role of Imported Cases and Favorable Meteorological Conditions in the Onset of Dengue Epidemics,” PLoS Negl Trop Dis, 2010; 4(8): e775.

[32] Liu-Helmersson J, Quam M, Wilder-Smith A, Stenlund H, Ebi K, Massad E, Rocklov J. “Climate Change and Aedes Vectors: 21st Century projections for dengue transmission in Europe,” EbioMedicine, 7 (2016): 267–277.

[33] Ebi K, Nealon J. “Dengue in a changing climate,” Environmental Research, 151 (2016): 115-123.

